# A Novel Integrated Nomogram for Predicting Prognosis in Pediatric Dilated Cardiomyopathy

**DOI:** 10.64898/2026.05.29.26354421

**Authors:** Ying Dai, Yan Wang, Youfei Fan, Hongjie Sun, Zhiying Dai, Ziding Tian, Peixing Wang, Hailin Jia, Li Zhang, Bo Han

## Abstract

**Background:** Pediatric dilated cardiomyopathy (DCM) is a leading cause of heart failure and transplantation, with variable prognosis and high early mortality. This study developed and validated a nomogram predicting short-term mortality risk to guide clinical decisions.

**Methods:** The data were sourced from the Pediatric Cardiomyopathy Database at Shandong Provincial Hospital. Cox regression analysis was conducted to determine outcome-associated factors, and a nomogram was developed to estimate 1, 3, and 5year mortality risks for children with DCM. Model effectiveness was assessed through the concordance index (C-index) and area under the receiver operating characteristic curve (AUC). Additionally, calibration curves and decision curve analysis (DCA) were employed to evaluate the model’s predictive accuracy and clinical relevance.

**Results:** A cohort of 106 children diagnosed with primary DCM and who underwent genetic analysis was studied, with a median diagnostic age of 10 months (ranging from 5 to 84 months), comprising 50 girls (47.2%). The rate of detecting genetic mutations was 28.3%, uncovering 14 gene variants linked to DCM, with *TTN* mutations being the most common. Both univariate and multivariate Cox regression analyses indicated that both sex and NT-proBNP levels had a significant impact on survival rates among pediatric DCM patients.The model exhibited strong discriminative performance, calibration, and clinical net benefit, as assessed by the C-index, calibration plots, and decision curve analysis (DCA).

**Conclusions:** The prediction model created in this research shows strong accuracy in forecasting survival rates at 1, 3, and 5 years for children with DCM, highlighting its significant relevance in clinical settings.

## Background

Dilated cardiomyopathy (DCM) is characterized by the enlargement of the left ventricle and impaired systolic function, with no identifiable hemodynamic reasons. Most individuals affected by this condition face worsening heart failure and its associated complications, positioning DCM as a primary contributor to heart failure and the need for cardiac transplants in pediatric populations. The condition tends to progress quickly, carries a significant risk of complications, and poses a notable threat of early death(1). Research shows that the yearly occurrence of pediatric DCM is around 0.5 to 0.7 cases per 100,000 children, with a one-year survival rate ranging from 69% to 72% post-diagnosis and a five-year survival rate between 54% and 63%. Mortality rates are particularly high shortly after the disease begins, especially among infants and young children, who also show higher incidence and poorer outcomes(2, 3). Despite improvements in heart failure medications, mechanical support technologies, and transplant management, the rate of survival without transplantation for children and adolescents with DCM, while better than in previous years, is still lacking. Additionally, long-term results can differ widely based on the underlying cause, the age at which the disease starts, and the specific structural and functional characteristics of the heart muscle(4). Some patients, however, experience significant recovery of heart function and reverse remodeling, especially when the underlying causes are reversible (like certain myocarditis cases, conditions related to load, or specific genetic factors with low penetrance) and when they receive prompt, standardized treatment. This situation calls for careful risk assessment in clinical settings to determine whether to adopt a \watchful waiting\ approach for recovery or to proceed with referrals for mechanical support or transplantation(5, 6).

While current biomarkers like LVEF, NYHA classification, and CPET, as well as general heart failure scoring systems, play crucial roles in evaluating the prognosis of DCM, their effectiveness and practical application are hindered by issues such as inadequate measurement reliability, varying outcome metrics, and the absence of a comprehensive approach that includes genetic, myocardial structural, and electrophysiological data(7, 8). There is a pressing demand among healthcare professionals for an accessible tool that can quickly assess individual risk based on a limited set of commonly used clinical variables, facilitating more accurate personalized risk assessment and intervention criteria.

A nomogram is a dynamic scoring framework developed from various essential factors that presents complex data visually as an accessible statistical instrument. This tool facilitates accurate assessment of personalized risk probabilities for particular outcomes, such as disease advancement or mortality, for individual patients(9). Nomograms are extensively utilized in fields like oncology for tailored predictions regarding postoperative recurrence, recurrence-free survival, and overall survival rates. They are also employed in cardiovascular medicine to evaluate lifetime risks associated with atherosclerosis and to stratify events in conditions like structural heart disease and cardiomyopathy, including the risk of sudden death in hypertrophic cardiomyopathy. Additionally, these tools are valuable in surgical and perioperative settings, aiding in crucial decision-making processes such as selecting treatment plans, determining follow-up intervals, and managing resource distribution(10-13).

The objective of this research was to develop nomograms that forecast mortality rates at 1, 3, and 5 years for pediatric patients diagnosed with dilated cardiomyopathy, thereby enhancing the accuracy of survival predictions and aiding in clinical decision-making.

## Methods

### Data Source

A total of 106 children diagnosed with primary dilated cardiomyopathy (DCM) were included in this research, all of whom received genetic testing. These patients were treated in the Pediatric Cardiology and Pediatric Intensive Care Unit at Shandong Provincial Hospital from January 2004 to January 2025. The diagnosis of primary DCM was confirmed during the initial clinical assessment at the hospital, with diagnostic and treatment plans developed collaboratively by a minimum of two specialists in pediatric cardiology.

### Inclusion and Exclusion Criteria

The 2019 AHA Scientific Statement on the Classification and Diagnosis of Cardiomyopathy in Children defines dilated cardiomyopathy (DCM) as a condition characterized by the enlargement of the left ventricle and impaired systolic function, which cannot be attributed to hemodynamic issues(4). If left ventricular dilation and systolic dysfunction continue despite suitable interventions due to underlying pathophysiological or anatomical reasons, they are also categorized as DCM.

The criteria for participation in this research were established as: (1) fulfillment of the previously mentioned diagnostic standards, where left ventricular enlargement is characterized by a left ventricular end-diastolic dimension or volume that surpasses 2 Z-scores above the average for the population, taking into account body size, sex, and/or age; and left ventricular overall systolic impairment indicated by an LVEF of less than 50%; (2) participants must be 18 years old or younger.

Individuals were not included in the study if they met any of these conditions: (1) age over 18; (2) existence of secondary factors contributing to heart enlargement and reduced systolic function, including physiological issues (like sepsis), unusual loading scenarios (such as aortic coarctation), or myocardial ischemia (for instance, an abnormal origin of coronary arteries); (3) data gaps greater than 50%; (4) instances where the ultimate diagnosis post-admission ruled out primary dilated cardiomyopathy.

### Genetic Testing Methods

Consent was secured from the guardians of all participants, who each completed an informed consent document. Genomic DNA was isolated from the patients’ peripheral blood samples and analyzed using next-generation sequencing (NGS). Whole-exome sequencing (WES) was chosen for the genetic analysis, primarily carried out by Fujun Gene Testing Company. Following this, Sanger sequencing was performed on both the patients and their family members to trace the mutations’ origins and evaluate the genetic testing outcomes. The Ethics Committee of Shandong Provincial Hospital approved this study, which was executed in full compliance with the Declaration of Helsinki established by the World Medical Association.

### Data Collection

During the first appointment, we recorded the fundamental characteristics of the patients, which encompassed the child’s sex, age at the time of diagnosis, weight, family history of disease, clinical manifestations (including syncope, fatigue, palpitations, etc.), growth failure, previous transient ischemic attacks or strokes, and any existing congenital heart conditions (such as patent foramen ovale [PFO] or atrial septal defect [ASD]). We also noted the cardiac functional classification and any instances of resuscitated ventricular fibrillation or cardiac arrest. For infants aged one year or younger, we utilized the modified Ross score for evaluation [Class I (0-2 points), Class II (3-6 points), Class III (7-9 points), or Class IV (10-12 points)], while for those older than one year, we assessed cardiac function using the New York Heart Association (NYHA) functional classification (12-item scale). Laboratory tests and cardiac evaluations included measurements of N-terminal pro-brain natriuretic peptide (NT-proBNP), serum potassium levels (normal range 3.5-5.0 mmol/L), alanine aminotransferase (ALT) (normal range 0-50 U/L), and hemoglobin (Hb), all obtained from blood samples taken on the day of admission and reviewed through the electronic medical record system. The electrocardiogram findings recorded included the presence of ventricular arrhythmias, QT interval prolongation, conduction blocks, and elevated left ventricular voltage. Each patient’s weight was documented, and body surface area (BSA) was calculated. An echocardiographic assessment was conducted, which included the evaluation of left ventricular ejection fraction (LVEF) and left ventricular end-diastolic dimension (LVEDD). We accounted for age and body surface area effects by calculating the Z-score for LVEDD.

All patient management strategies were thoroughly recorded. The treatment of heart failure involved various medications, mainly focusing on angiotensin-converting enzyme inhibitors (ACEIs) and beta-blockers. Amiodarone was used as the primary treatment for severe tachyarrhythmias. In cases of critically ill patients, positive inotropic support was offered, utilizing options such as oral digoxin or intravenous dopamine, dobutamine, or milrinone. Those with thrombus within the heart were treated with anticoagulants.

### Data Analysis

Continuous variables were summarized as median values along with interquartile ranges (IQR) and compared across groups utilizing the Mann-Whitney U test. Categorical data were represented as counts and percentages, analyzed through either the chi-square test or Fisher’s exact test, depending on the context. Initially, a univariate Cox regression analysis was conducted to pinpoint potential factors linked to negative outcomes in dilated cardiomyopathy. Variables that yielded a *P*-value of less than 0.05 in the univariate analysis were then included in a multivariate Cox regression model to identify independent predictors with a significance level of *P* < 0.05. Nomograms for predicting mortality at 1, 3, and 5 years in patients with dilated cardiomyopathy were developed using the RMS package in R. The model’s discriminative performance was evaluated through Harrell’s concordance index (C-index) and the area under the receiver operating characteristic (ROC) curve. Calibration curves were created using the Bootstrap method with 1000 resamples to assess the model’s calibration accuracy. Lastly, decision curve analysis (DCA) was performed to evaluate the model’s clinical applicability.

Statistical evaluations were conducted utilizing R version 4.3.3, developed by the R Foundation for Statistical Computing in Vienna, Austria. A *P* value of less than 0.05 on both sides was deemed to indicate statistical significance.

## Results

### Patient Characteristics

A cohort of 106 children diagnosed with primary dilated cardiomyopathy (DCM) was studied, with a median age of 10 months at the time of diagnosis (ranging from 5 to 84 months). This group included 56 boys, accounting for 52.83% of the participants. Genetic analysis indicated a positivity rate of 28.30%, identifying mutations across 14 different genes, which encompassed those related to sarcomeres (such as *TTN* and *MYH7*), mitochondrial function, cytoskeletal components, and transcriptional regulation (see Figure 5). The average follow-up period lasted 31.50 months (with a range of 9.25 to 68 months), during which 17 participants (around 16%) passed away. Among the survivors, 24 individuals (26.97%) tested positive for genetic mutations, while 6 individuals (35.29%) from the deceased group were also found to be genetically positive.

In contrast to the survival cohort, the group of individuals who passed away exhibited markedly elevated levels of NT-proBNP (18.55 ng/mL [12.71–34.07] compared to 5.52 [1.95–15.00], *P* = 0.002), a reduced rate of digoxin administration (58.82% versus 85.39%, *P* = 0.018), and a shorter follow-up period (3.00 months [1.50–6.00] against 46.00 months [17.00–78.00], *P* < 0.001). Echocardiographic assessments revealed that the left ventricular end-diastolic diameter (LVEDD) was 4.37 (3.70–5.04) cm in the survival group and 4.44 (3.71–6.00) cm in the deceased group, while the left ventricular ejection fraction (LVEF) was recorded at 30.00 (25.00–39.00)% for both groups. Overall, the imaging parameters, laboratory results, medication usage, and complications were largely comparable between the two groups, with no notable differences identified.

### Variable Analysis and Selection

Single-variable analysis indicated that three key factors were linked to unfavorable outcomes in individuals with dilated cardiomyopathy (see Table 1). These factors— NT-proBNP levels, gender, and the use of digoxin—were recognized as possible indicators and were later incorporated into the multivariate Cox regression analysis (refer to Table 2). The multivariate analysis revealed that NT-proBNP (hazard ratio [HR] = 1.043, 95% confidence interval [CI] = 1.014–1.074, *P* = 0.004), gender (HR = 2.704, *P* = 0.091), and digoxin administration (HR = 0.281, *P* = 0.015) were correlated with patient outcomes.

### Construction of the Nomogram

A prediction model in the form of a nomogram was developed utilizing the rms package, which included key variables determined through multivariate Cox regression analysis (refer to Table 3). This model aims to estimate survival rates at 1, 3, and 5 years for the patient cohort (see Figure 1).

**Figure 1.**
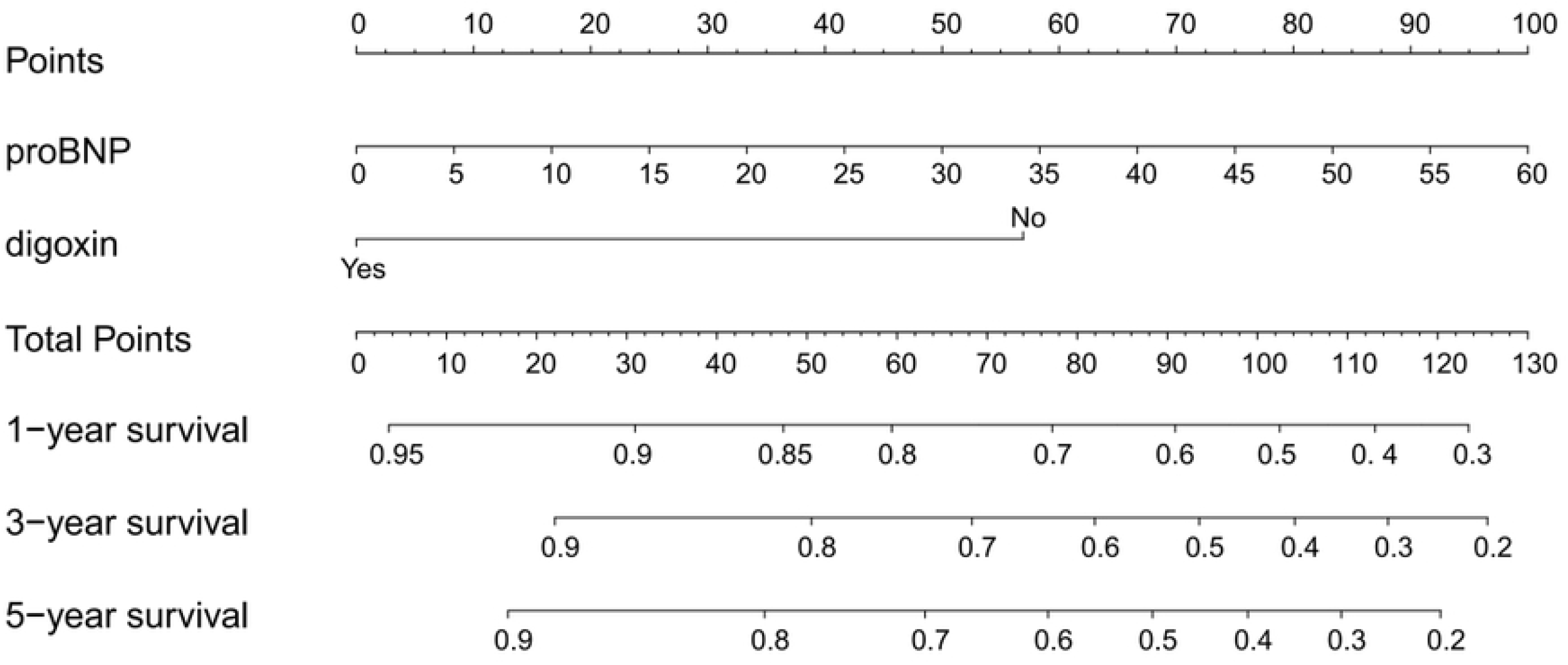
Nomogram for predicting the risk of death at 1, 3, and 5 years.

### Performance Evaluation of the Nomogram

The analysis of the ROC curve revealed that the model performed well in forecasting survival across various follow-up intervals, achieving an AUC of 0.770 for predicting 1-year survival, along with AUCs of 0.749 and 0.753 for 3-year and 5-year survival predictions, respectively (see Figure 2). The calibration curves generated at these time points closely matched the ideal 45-degree diagonal line, signifying a strong correlation between the predicted outcomes and the actual results (refer to Figure 3). Additionally, the decision curve analysis (DCA), which illustrates net benefit, validated the model’s effective clinical application in mortality prediction (illustrated in Figure 4).

**Figure 2.**
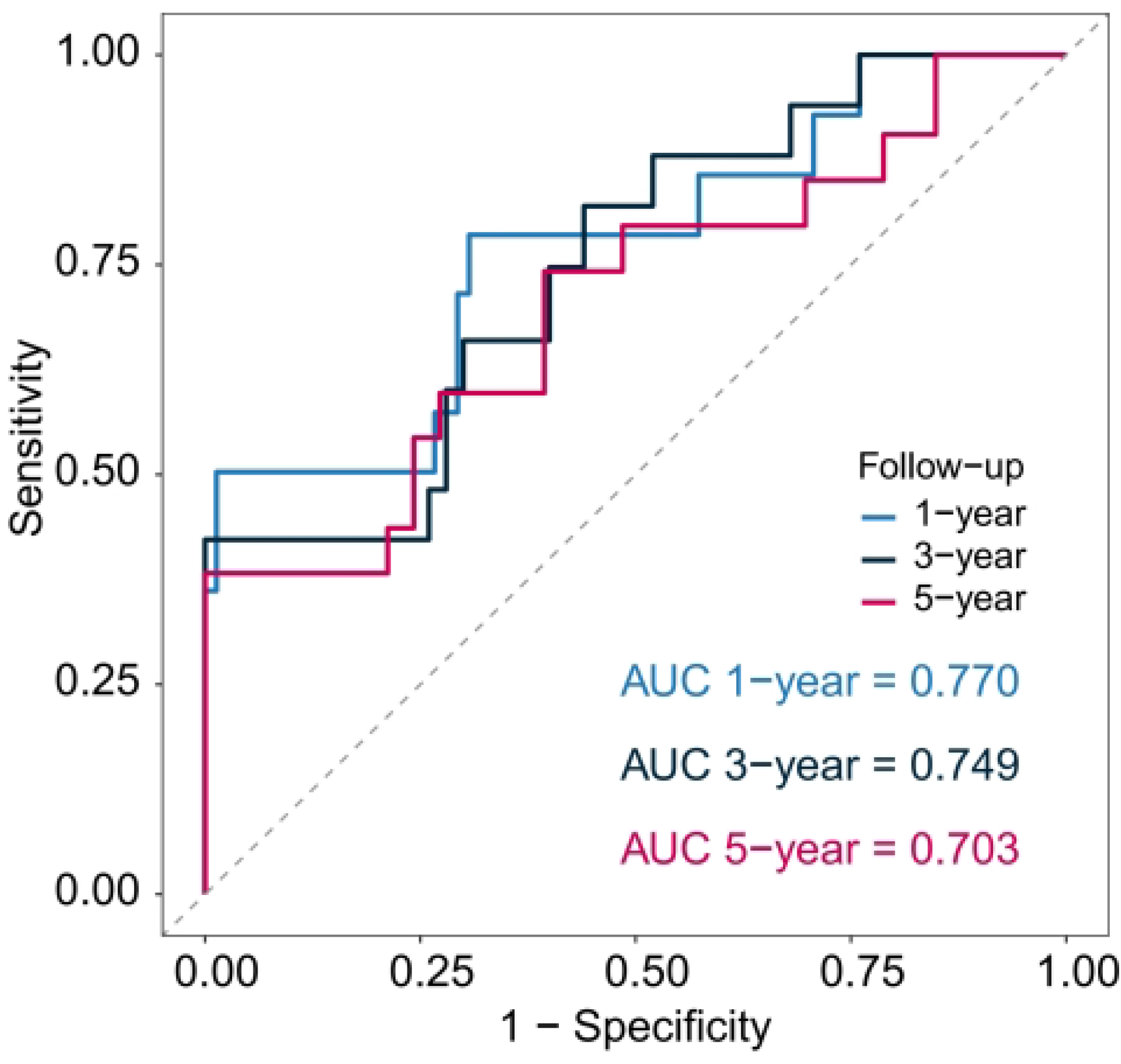
Time-dependent ROC curves of the risk prediction model for death at 1, 3, and 5 years.

**Figure 3.**
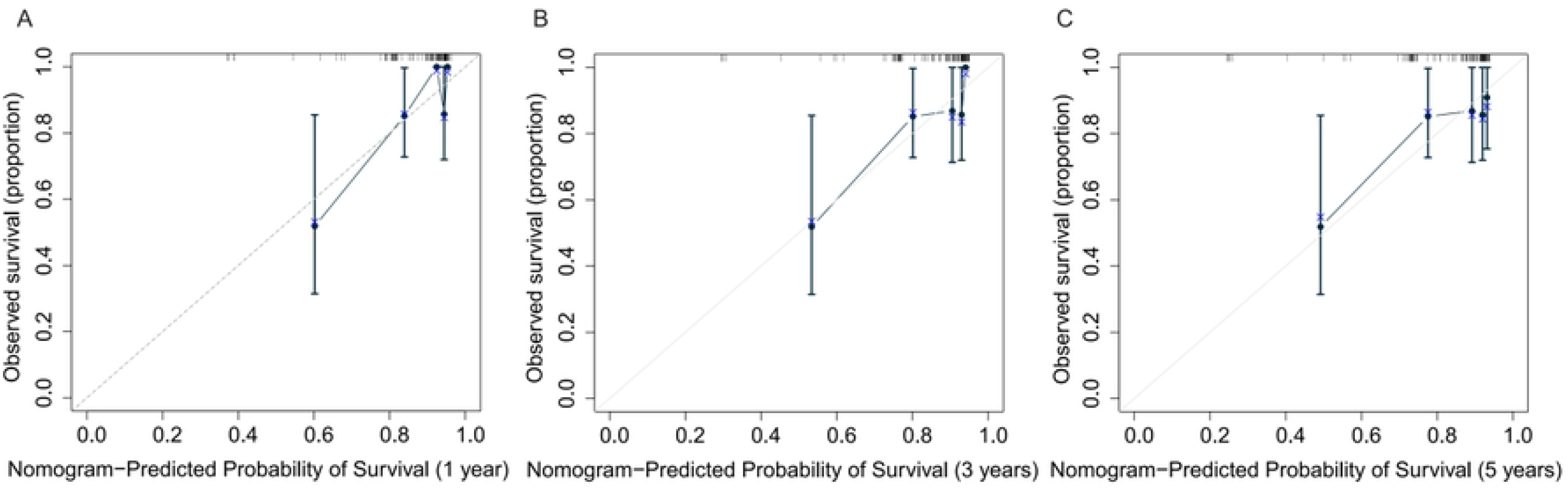
Calibration curves of the risk prediction model for death at 1 (A), 3 (B), and 5 (C) years.

**Figure 4.**
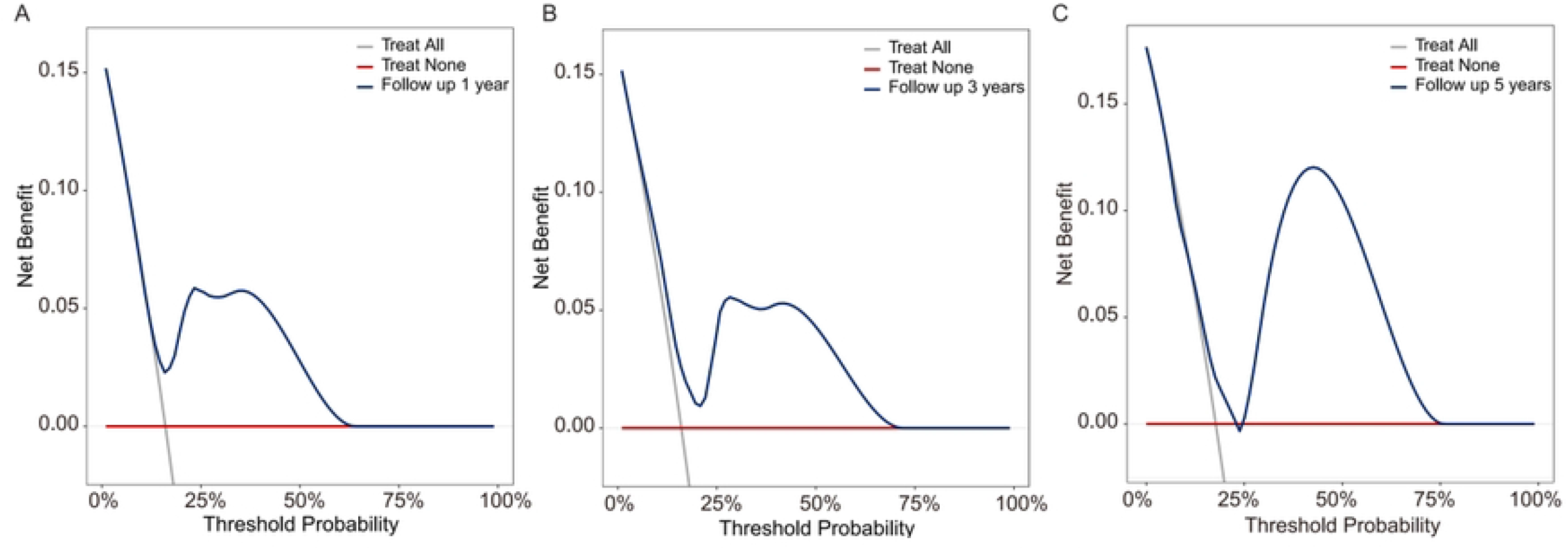
Decision curves of the risk prediction model for death at 1 (A), 3 (B), and 5 (C) years.

**Figure 5.**
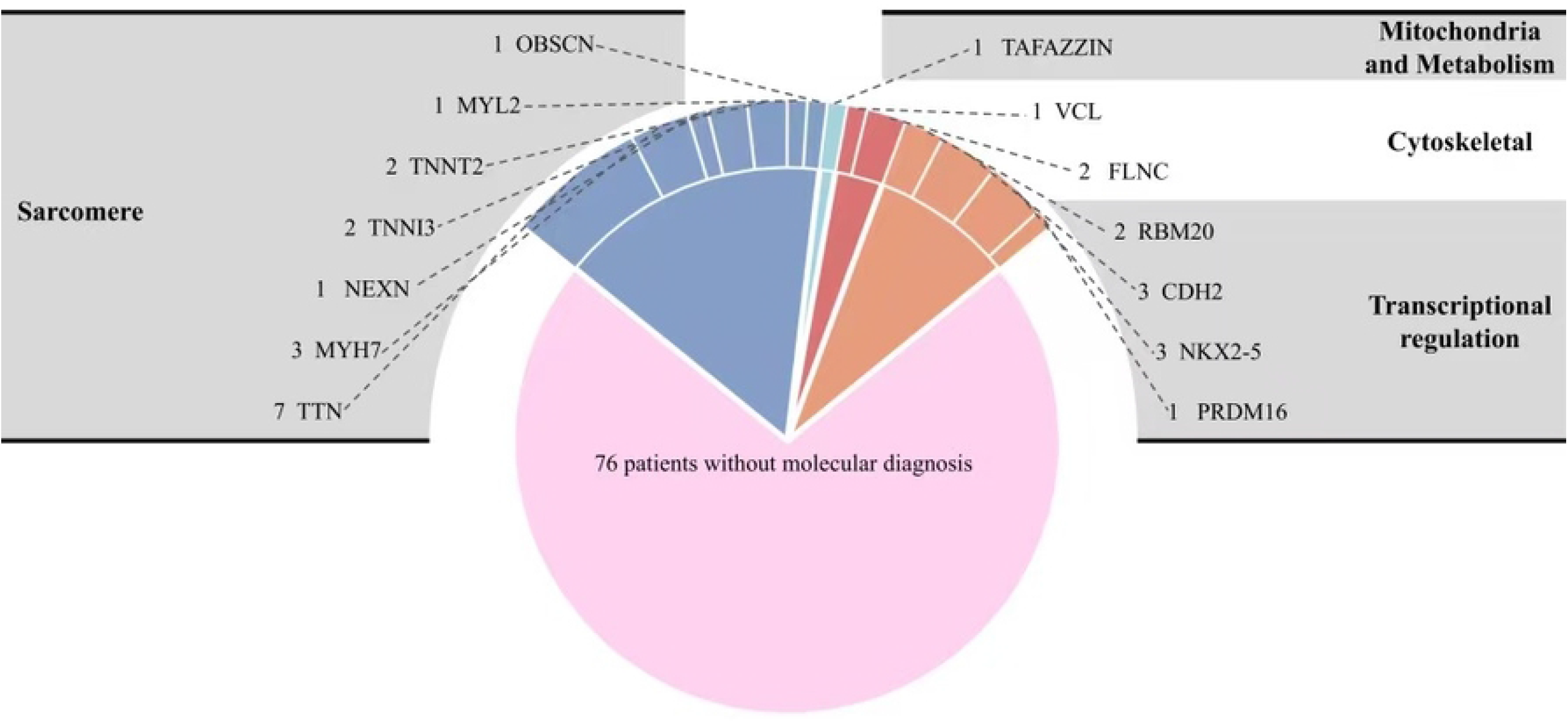
Distribution map of pathogenic or potentially pathogenic gene mutations in primary DCM in children.

## Discussion

Dilated cardiomyopathy (DCM) is a serious heart condition that significantly contributes to heart failure and the need for heart transplants, as well as being a major cause of sudden cardiac death in those under 35 years old(14, 15). This disease is marked by the enlargement of the left or both ventricles and impaired systolic function, posing a considerable risk to health, with only a 50-60% survival rate over five years(1, 2). In children, DCM is the most prevalent form of cardiomyopathy, occurring at a rate of about 0.57 cases per 100,000 annually, and has a five-year survival rate of roughly 70-75%, making it the primary reason for heart transplants in this age group(3). Pediatric DCM shows significant genetic diversity, with mutations in various genes linked to its onset, affecting heart function through different genetic mechanisms and leading to a range of clinical presentations(16, 17). The high rates of incidence, progression of heart failure, and mortality, along with the considerable financial strain on families, have heightened the attention of healthcare professionals and researchers on treatment options for children with DCM. Despite challenges related to clinician experience and patient adherence, several scoring systems, including the Pediatric Cardiomyopathy Registry (PCMR) criteria, Ross heart failure classification, New York University Pediatric Heart Failure Index (NYU-PHFI), Pediatric Heart Failure Score, and Seattle Heart Failure Model, have been utilized to evaluate prognosis in pediatric DCM, though results may not always align with actual clinical outcomes. At present, there is a notable absence of validated tools for risk stratification to forecast mortality over time in young patients with DCM. Consequently, there is a pressing need for the creation of a more user-friendly and consistent predictive model that can evaluate both prognosis and transplant-free survival across various time intervals in pediatric DCM patients.

This research conducted a longitudinal follow-up of children diagnosed with primary DCM, uncovering that the group of deceased individuals showed markedly higher levels of biomarkers and lower rates of digoxin usage. The multivariate Cox regression analysis pinpointed NT-proBNP, gender, and digoxin administration as independent prognostic indicators. A nomogram developed from these variables exhibited strong capabilities in distinguishing and accurately predicting survival outcomes. Validation through ROC curves, calibration curves, and decision curve analyses confirmed the model’s effectiveness as a decision-making resource for risk assessment and personalized treatment strategies for children with DCM.

Recent advancements in genetic sequencing technologies have led to significant developments in the genetic analysis of dilated cardiomyopathy (DCM). To uncover new pathogenic genes and mutations, researchers have utilized various strategies for the swift and accurate interpretation of harmful variants in pediatric DCM cases. Techniques like whole exome sequencing (WES) and whole genome sequencing (WGS) are effective in quickly detecting rare pathogenic variants, with WES being the most commonly used diagnostic approach for hereditary DCM(18). Moreover, conventional techniques such as familial linkage analysis remain crucial for identifying pathogenic genes in familial cases of DCM. We gathered extensive data on pediatric patients from the pediatric cardiomyopathy database at Shandong Provincial Hospital, which includes thorough clinical details of children diagnosed with DCM at our facility. All patients underwent WES, and Sanger sequencing was conducted on both the patients and their family members to trace the mutation origins, allowing for a deeper analysis of the genetic testing outcomes in pediatric DCM. The data collected were intended to create a computational model that predicts adverse outcomes at 1, 3, and 5 years post-DCM diagnosis. The model incorporated objective clinical parameters that are routinely gathered in standard pediatric cardiology practice. Following this, we performed a thorough evaluation and validation of the model using metrics such as the concordance index (C-index), area under the receiver operating characteristic curve (AUC), calibration curves, and decision curve analysis (DCA) to measure its discriminative power, calibration accuracy, and clinical relevance.

Pediatric dilated cardiomyopathy (DCM) shows considerable genetic diversity, featuring many rare genetic variants, most of which are classified as variants of uncertain significance (VUS)(19).This complexity requires thorough clinical phenotyping, analysis of familial co-segregation, and functional validation for accurate interpretation. In our research, around 28.3% of pediatric DCM patients were found to have either definitive pathogenic (P) or likely pathogenic (LP) mutations, a figure that is consistent with earlier estimates of genetic causes in DCM, which range from 30% to 50%(20). We discovered 14 unique genetic mutations, primarily located in genes associated with sarcomere proteins (such as *TTN, MYH7, TNNI3, TNNT2*) and those involved in transcriptional regulation (including *NKX2-5, CDH2, RBM20*). The *TTN* gene mutations were the most prevalent in our sample, aligning with previous findings that highlight *TTN* mutations as the leading cause of DCM(21). Additionally, mutations in sarcomeric genes like *MYH7, TNNT2*, and *TNNI3* were frequently observed, emphasizing the critical importance of these genes in DCM development(22-24). Variants in non-sarcomeric genes also deserve significant consideration; we recorded nine instances of mutations in transcriptional regulatory genes (three each in *NKX2-5* and *CDH2*, two in *RBM20*, and one in *PRDM16*), three cases involving cytoskeletal genes (two in *FLNC* and one in *VCL*), and one case with a *TAFAZZIN* mutation linked to mitochondrial function. This genetic variability illustrates the intricate genetic framework of pediatric DCM, where specific mutations may be associated with varying clinical presentations and outcomes. Genetic profiling can aid in identifying DCM patients with different prognostic implications, thus laying the groundwork for precise diagnoses and tailored treatment approaches.

Pediatric dilated cardiomyopathy (DCM) is a complex condition influenced by various risk factors that affect patient outcomes. Analysis using multivariable regression indicated that factors such as sex, NT-proBNP levels, and the use of digoxin significantly impact survival rates among stroke patients. In the pediatric demographic, DCM shows notable differences based on gender. Specifically, being male has been recognized as a distinct risk factor for poorer outcomes in children with DCM. Epidemiological research indicates that boys with this condition tend to experience faster heart failure progression and a greater need for heart transplants compared to girls(3, 25, 26). This gender difference may stem from hormonal variations, where the protective effects of estrogen are less effective in prepubescent females, while males are more affected by testosterone-related inflammatory responses and heart remodeling processes that accelerate disease advancement(27, 28). Prior studies have established gender as a key independent predictor of outcomes. It has been observed that male patients often present with more severe left ventricular dysfunction and reduced ejection fractions at the time of diagnosis(25). This gender-based prognostic difference may be linked to hormonal fluctuations, variations in gene expression, and differing immune responses to heart injury(29, 30). Additionally, specific genetic mutations, such as those in the DMD gene leading to Duchenne muscular dystrophy-associated cardiomyopathy, predominantly impact males, further intensifying the prognostic disparities between genders(31).

In addition, high levels of NT-proBNP are strongly linked to the severity of pediatric dilated cardiomyopathy. The processes of ventricular remodeling and stress on cardiomyocytes lead to increased production and release of the brain natriuretic peptide precursor, which significantly raises the levels of NT-proBNP in circulation, indicating the extent of cardiac function decline(32, 33). Research has shown that NT-proBNP is not only a crucial marker for evaluating the prognosis of pediatric dilated cardiomyopathy, but its initial levels and fluctuations can also forecast the likelihood of cardiovascular incidents and survival outcomes in affected children(34, 35).

Dilated cardiomyopathy (DCM) stands as the primary contributor to heart failure in children, necessitating a thoughtful approach to treatment options that influence the long-term survival of these young patients. Additionally, the use of digoxin may enhance outcomes for children suffering from DCM, as it has been shown to lower the risk of cardiovascular-related deaths by improving heart muscle function and regulating heart rate. While this advantage is well-documented in adult heart failure cases, conclusive data in the pediatric and adolescent demographics is still insufficient(36, 37). This study found that digoxin serves as an independent protective factor for children with dilated cardiomyopathy, underscoring its significant role in managing heart failure in this population.

The conventional evaluation of prognosis in pediatric dilated cardiomyopathy (DCM) has largely depended on the clinical judgment of physicians and isolated metrics, such as left ventricular ejection fraction and NT-proBNP levels, which do not offer a comprehensive or personalized predictive approach. This research introduces a nomogram model that amalgamates various objective metrics, presenting a straightforward and intuitive visualization tool for personalized risk evaluation and ongoing management. This advancement aids in accurately stratifying risk among pediatric patients, pinpointing high-risk groups, and enhancing treatment strategies for healthcare providers. Notably, this study marks the inaugural development of a prognostic nomogram tailored specifically for the pediatric DCM demographic, addressing the absence of quantitative predictive instruments in pediatric dilated cardiomyopathy. Its predictive accuracy significantly exceeds that of traditional techniques, offering new scientific insights for precision medicine in this area.

This research has a number of constraints. To begin with, the retrospective nature of the study conducted at a single center could have led to selection bias. Additionally, we omitted specific echocardiographic metrics and emerging cardiac biomarkers that could provide prognostic insights but are not typically assessed in children. Lastly, the absence of validation in an independent cohort restricts our capacity to evaluate the effectiveness of this nomogram in various pediatric groups. Consequently, there is a need for multicenter prospective research and external validation to establish the clinical relevance and reliability of this nomogram.

## Conclusion

Utilizing objective demographic data and clinical metrics, this research developed an accessible nomogram to predict the mortality risk at 1, 3, and 5 years for children diagnosed with dilated cardiomyopathy after the onset of the condition. This tool aids healthcare professionals in conducting accurate risk evaluations and creating tailored treatment plans, while also empowering families to make well-informed healthcare choices. However, our results necessitate additional validation and enhancement through multicenter studies involving larger participant groups.

## Data Availability

The data that support the findings of this study are available from the corresponding author upon reasonable request.

## Abbreviations

C-index: Harrell’s concordance index
IQR: Interquartile-range
AUC: The area under the curve
DCA: Decision curve analysis
HR: Hazard ratio
ROC: Receiver operating characteristics curve.

